# Trajectory and genetic correlation mapping of cross-disease risks following incident type 2 inflammatory diseases

**DOI:** 10.1101/2025.09.02.25334921

**Authors:** Yask Gupta, Henning Olbrich, Diamant Thaci, Philip Curman, Ralf J. Ludwig

## Abstract

Type 2 inflammatory diseases (T2IDs) such as atopic dermatitis (AD), asthma, food allergy, eosinophilic esophagitis (EoE), chronic sinusitis (CS), and others, are immune-mediated conditions that frequently co-occur across organ systems and contribute to substantial disease burden. While the atopic march has long served as a conceptual model for T2ID progression, it remains unclear whether this linear sequence captures the broader comorbidity structure of T2IDs. In this study, we systematically analyzed comorbidity patterns across nine T2IDs using two complementary methodological approaches: Large-scale real-world data (RWD) from over 129 million electronic health records and genome-wide genetic correlation, including data from three novel (asthma, chronic sinusitis and allergic rhinitis) metanalyses of genome-wide association studies. Retrospective matched cohort analyses revealed a highly interconnected disease network in which CS emerged as a central driver of downstream comorbidities, increasing the risk of multiple other T2IDs across diverse patient populations. AD, food allergy, and vasomotor and allergic rhinitis also showed strong cross-disease effects. Subgroup analyses uncovered notable age-, sex-, and race-specific disparities. Genetic correlation analyses supported shared heritability among many disease pairs and integration with RWD allowed to infer directionality. While some disease pairs showed discordant findings between genetic correlation and RWD, most associations aligned across both methodologies. Together, our results highlight that T2ID progression extends beyond the atopic march and is shaped by a complex and heterogeneous network of interactions. This work provides a data-driven framework to guide risk stratification, inform early preventive strategies, and prioritize upstream disease targets within the type 2 inflammatory spectrum.

## Introduction

Type 2 inflammatory diseases (T2IDs) represent a group of chronic, immune-mediated conditions affecting the skin, airways, and gastrointestinal tract. Despite their phenotypic diversity, T2IDs such as atopic dermatitis (AD), asthma, prurigo nodularis (PN), food allergy, eosinophilic esophagitis (EoE), chronic sinusitis (CS), vasomotor and allergic rhinitis (VMR/AR), chronic obstructive pulmonary disease (COPD), and bullous pemphigoid (BP) are united by overlapping inflammatory signatures, including the involvement of type 2 cytokines (interleukin [IL]-4, IL-5, IL-13), eosinophils, and IgE-driven pathways (1,2).

T2IDs are highly prevalent and exert a substantial burden on patients and healthcare systems. Given the high degree of comorbidity among T2IDs, the cumulative prevalence of having at least one of these diseases in the general population ranges between 30% and 50% (3,4). Although precise estimates of disability-adjusted life years (DALYs) and direct annual healthcare costs are not available for each of the mentioned T2IDs individually, available data suggest that DALYs range from 123 per 100,000 population (AD) to 926 per 100,000 population in COPD. In the United States alone, direct annual healthcare costs attributable to T2IDs are estimated between $5 billion and $31 billion, underscoring the substantial socioeconomic impact of these conditions (5–9).

Beyond their individual burden, T2IDs frequently co-occur, often across organ systems (10). A well-known example for this comorbidity pattern is the atopic march, in which early-onset atopic dermatitis is frequently followed by food allergy, AR, and asthma, highlighting a temporal and mechanistic progression across barrier organs (11,12). The concept is supported by longitudinal birth cohort studies such as the German Multicenter Allergy Study (MAS), which demonstrates sequential disease onset and provide mechanistic insights into skin barrier dysfunction, allergen sensitization, and Th2-driven inflammation (13). However, recent evidence has challenged the notion of a universal progression, i.e., the MAS noted that not all children with early-onset AD will subsequently develop asthma (14). Thus, not all patients follow the proposed trajectory in the atopic march, and alternative comorbidity patterns have emerged from population-based studies using data-driven clustering or latent class analysis. These findings suggest that the atopic march represents one of several possible trajectories within a broader and more heterogeneous network of T2IDs (15,16). However, this broader structure of T2ID comorbidity patterns across the full spectrum of T2IDs, and the extent to which these reflect shared genetic risk, remains incompletely understood. Furthermore, previous studies largely focused on single T2IDs, specific age groups, or limited outcome measures. In rare cases, disparities in sex or race were addressed.

In addition to clinical and epidemiologic evidence, genetic data are beginning to emerge that support shared susceptibility across T2IDs. Genome-wide genetic correlation analyses demonstrated heritability overlap for a subset of T2ID pairs, most notably between AD and asthma (17), or asthma and AR (18,19), where robust genetic correlations have been consistently reported. In some instances, Mendelian randomization (MR) studies demonstrated directional, potentially causal relationships, such as from AD to asthma (20). However, for the majority of T2ID combinations, especially those encompassing diseases such as EoE, PN, or BP, genetic correlation or data from MR studies remain limited or are entirely lacking. Overall, while initial evidence underscores the genetic interconnectedness of certain T2IDs, a comprehensive understanding of the genetic architecture underlying comorbidity across the full spectrum of T2IDs is still incomplete.

Taken together, several key knowledge gaps relating to T2ID trajectories remain: First, previous studies rarely examined comorbidity patterns across the full spectrum of T2IDs in a unified, systematic framework. Second, we are not aware of investigations have combined large-scale real-world data (RWD) with genetic correlation analyses to explore whether observed clinical associations reflect shared heritable risk. Third, although individual T2IDs have been associated with age-, sex-, and racial-disparities, it is unclear how these influence comorbidity trajectories across diseases. Addressing these gaps is essential for improving risk stratification and to guide the development of treatment decisions. To address these knowledge gaps, we conducted a comprehensive analysis of nine major T2IDs using retrospective cohort analyses from the TriNetX electronic health record (EHR) network, complemented by genome-wide genetic correlation estimates. This dual approach allowed us to (i) map cross-disease risks in a large, diverse populations, (ii) assess the degree of genetic overlap across T2IDs, (iii) provide direction of observed genetic correlations by integration with RWD, and (iv) compare cross-disease risks among T2IDs based on two independent methodologies, overall strengthening the results of our findings. From a clinical perspective, such insights could shape screening strategies for at-risk populations, support early intervention in disease trajectories, and help identify disease clusters that may benefit from shared therapeutic approaches.

## Methods

### Retrospective cohort studies

#### Ethics

The data reviewed is a secondary analysis of existing data, does not involve intervention or interaction with human subjects, and is de-identified per the de-identification standard defined in Section §164.514(a) of the HIPAA Privacy Rule. TriNetX is a federated network which facilitates collaboration between industry including pharmaceutical companies, research organizations, academia, and health care organizations (HCOs). TriNetX has a security and governance model that provides a federated platform of real-world-data (RWD, i.e., electronic health records (EHRs)) which is Health Insurance Portability and Accountability Act (HIPAA), General Data Protection Regulation (GDPR), and Lei Geral de Proteção de Dados (LGPD)-compliant. Thus, our study did not require Institutional Review Board approval.

#### Study design and database

A propensity-score matched (PSM) retrospective cohort study was conducted using the US Collaborative Network from the federated EHR database TriNetX, following established protocols (21,22). This network was selected due to the large number of documented EHRs and the high degree of covariate documentation (23). Patients diagnosed with an incident diagnosis of the following T2IDs were identified: AD, PN, BP, asthma, VMR/AR, CS, COPD, food allergy, and EoE. The risk for each T2ID to develop any of the other eight T2IDs was contrasted to that of non-T2ID controls, using PSM for better comparability. Three sensitivity analyses were performed to test the robustness of the findings. Sex-, race-, and age-stratified analyses were conducted to explore potential disparities relating to these factors. The analytical framework, definition of cohorts, matching factors, and outcomes were defined prior to data collection (**Figure 1**). The study was performed according to the STROBE guidelines (24).

**Figure 1.**
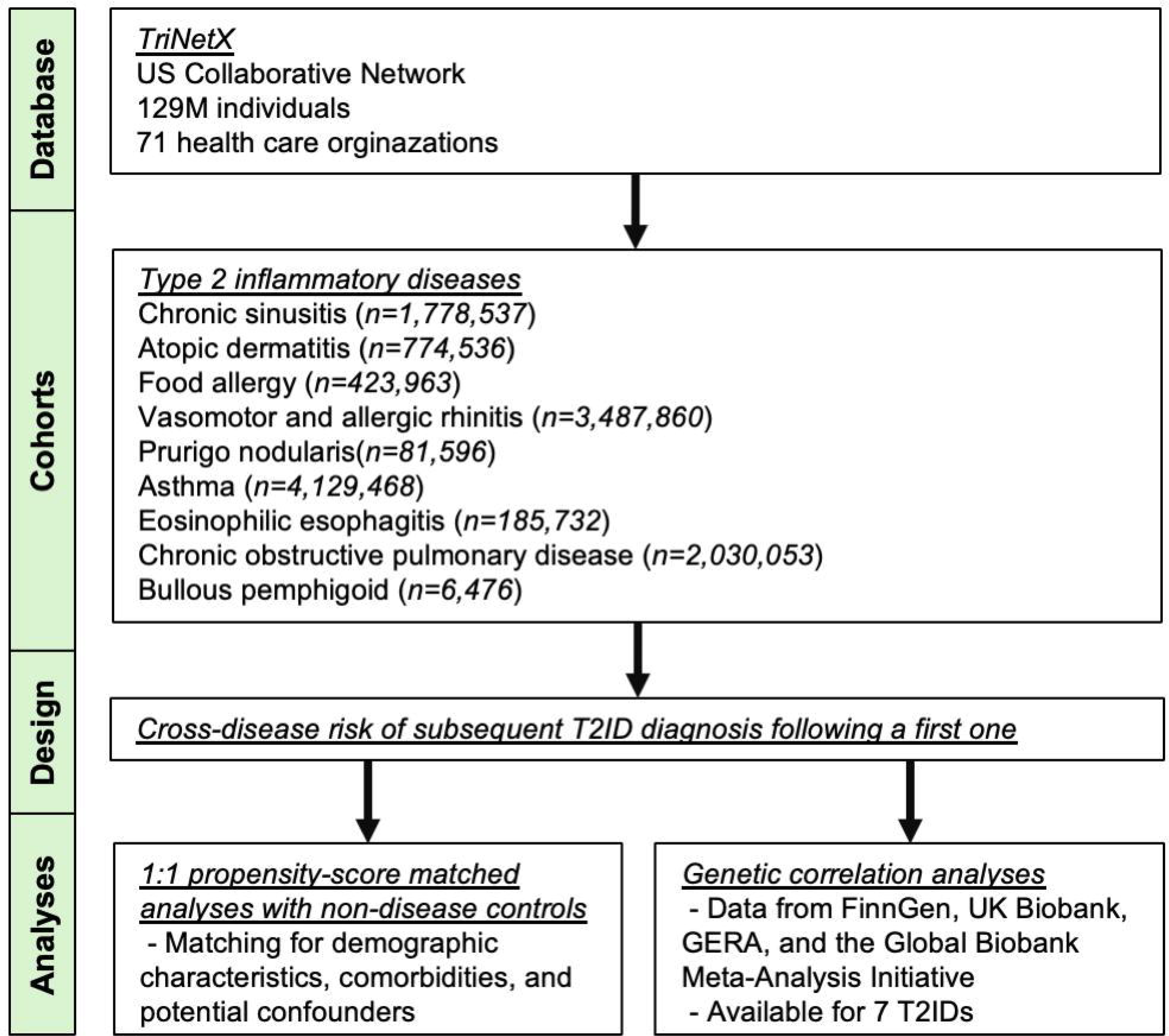
Flow chart of the study design and analytical framework.

#### Study population

The study was conducted from March to May 2025. At the time of analysis, access to over 129 million patients across 71 HCOs in the United States was provided. Only EHRs from patients with incident T2IDs and no prior history of any other T2ID were included. T2IDs were categorized into dermatological T2IDs (AD, PN, and BP), airway T2IDs (VMR/AR, CS, asthma, and COPD), and gastrointestinal T2IDs (EoE and food allergy). All diseases were defined by their respective International Classification of Diseases, 10^th^ Edition, Clinical Modification (ICD-10-CM) codes (**Supplement Table 1**). To exclude miscoding of BP for pemphigus or other pemphigoid diseases, we excluded all EHRs with a documentation of any other pemphigus or pemphigoid disease (**Supplement Table 1**). Non-T2ID controls were defined by Z00 (general healthcare encounter) and the absence of any T2ID prior to the healthcare visit. Index events were defined by the date of the incident T2ID diagnosis or, for controls, the date of the healthcare encounter.

#### Covariates

To minimize bias from potentially confounding variables, PSM was performed by establishing a covariate matrix that included demographic information, potential health hazards related to socioeconomic and psychosocial circumstances, and relevant concomitant diagnoses. Covariates included in all analyses were age at index (continuous), female sex (binary), self-reported White ethnicity (binary), potential health hazards related to socioeconomic and psychosocial circumstances (Z55-Z65, binary), overweight and obesity (E66, binary), and nicotine dependence (F17, binary). Matching was performed 1:1 using propensity-scores. Baseline characteristics were reported before and after matching (**Table 1**).

**Table 1.**
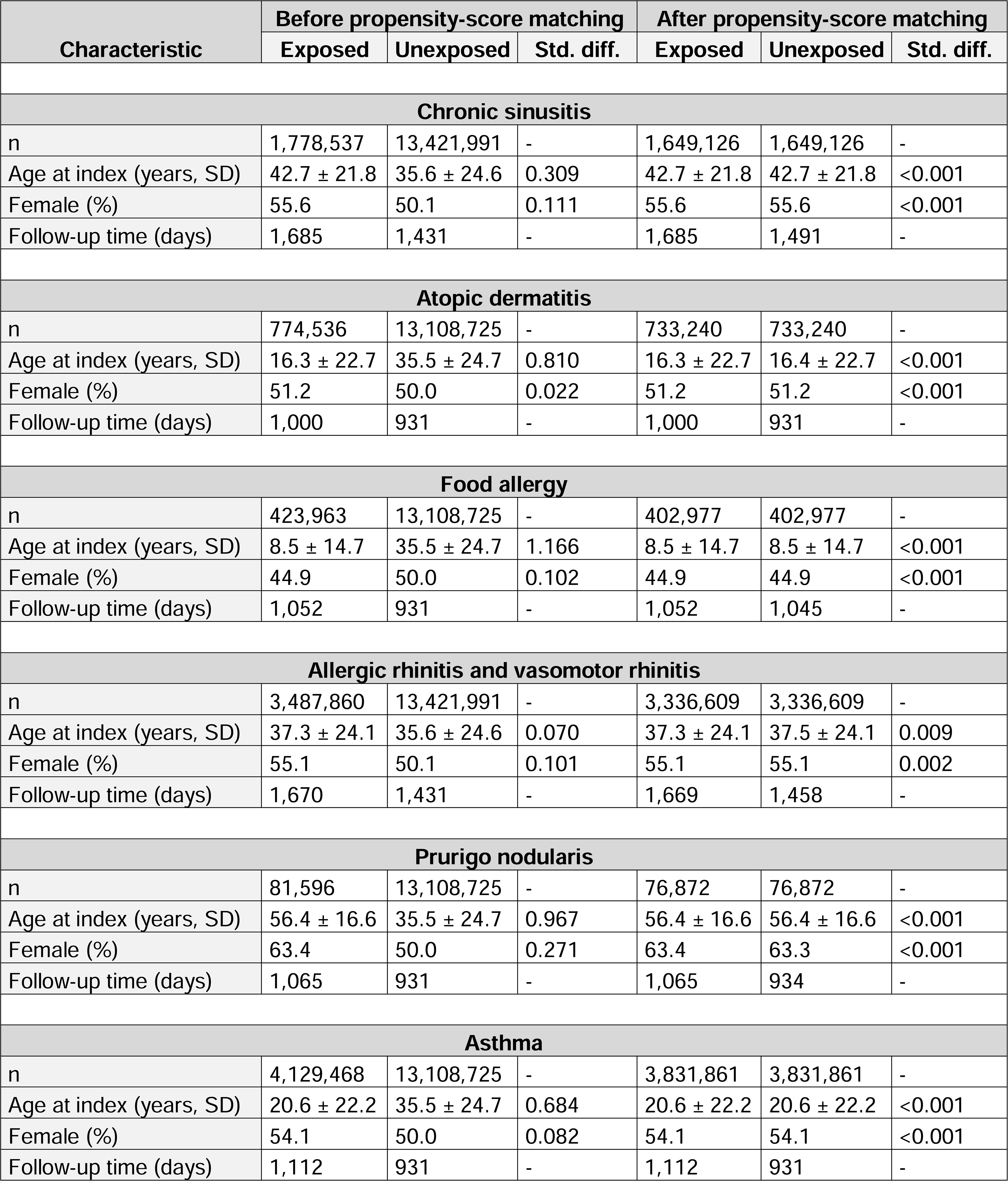
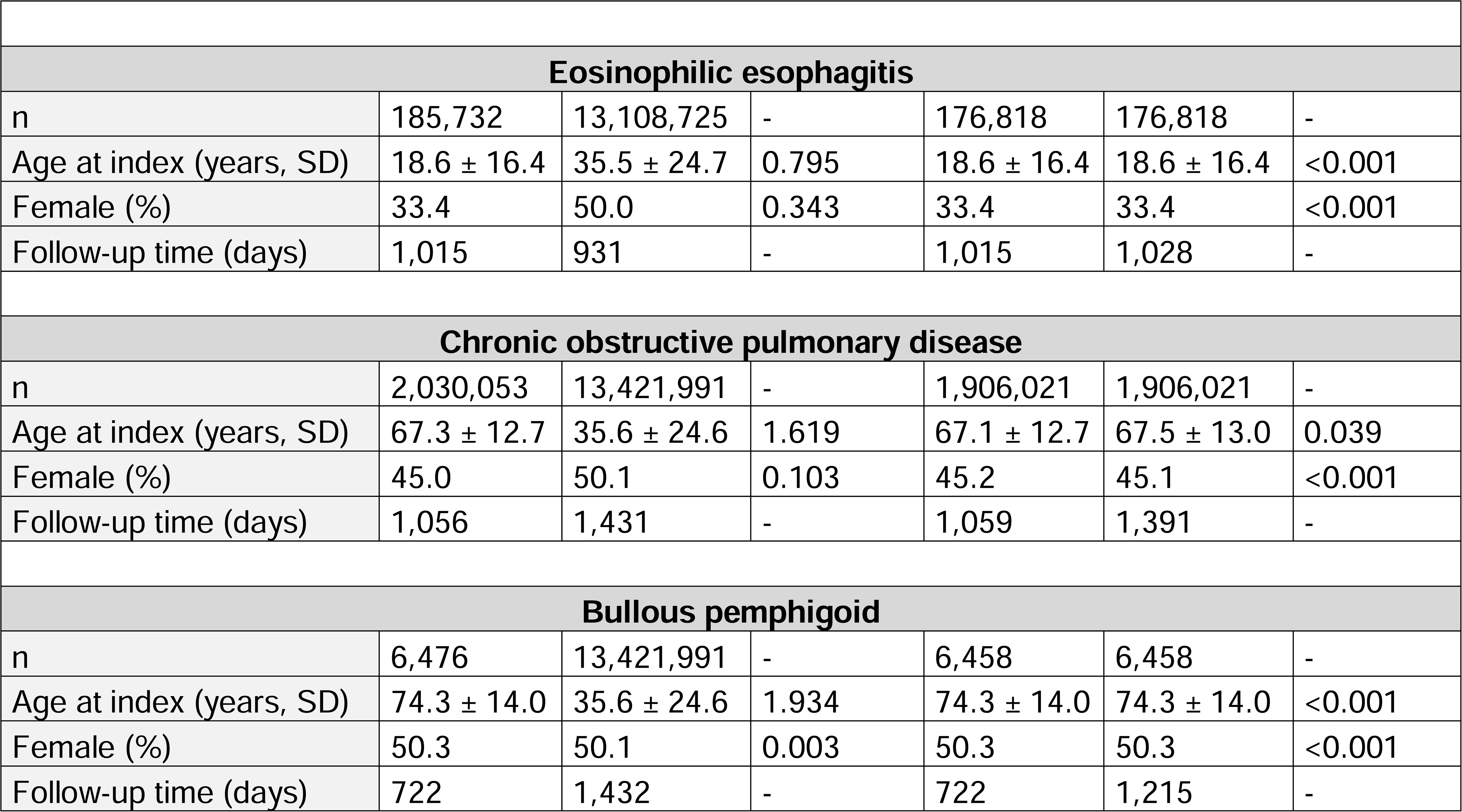
Summarized baseline characteristics for all study cohorts including cohort sizes, age at index, sex, and follow-up times for the primary analysis. *Abbreviations: SD: standard deviation; Std. diff.; standardized difference*

**Table 2.**
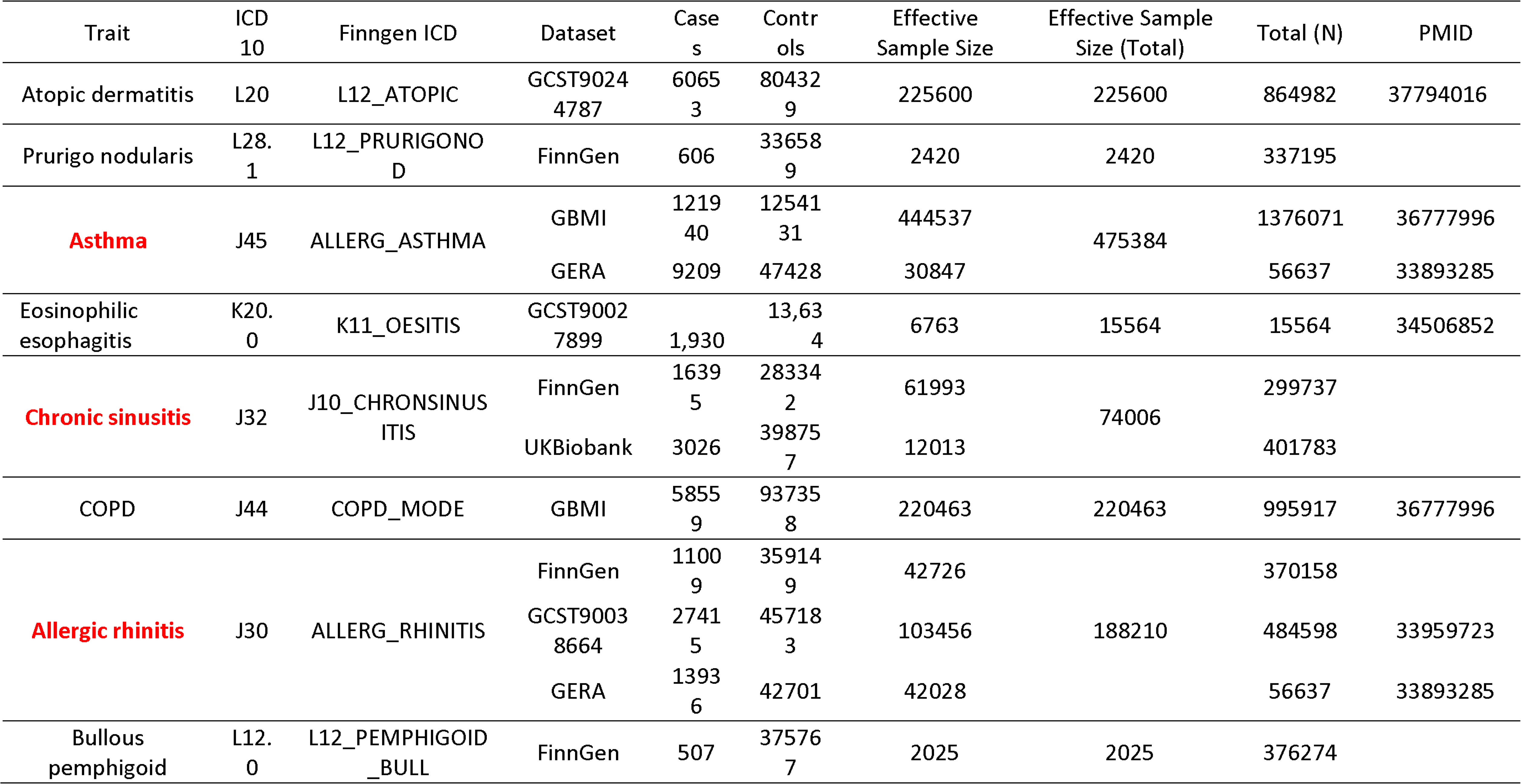
Sources of data for genetic correlational analysis. New metanalysis are depicted in red color. Summary statistics from non-FinnGen datasets are available upon request. FinnGen-derived summary statistics were used under data use restrictions and cannot be publicly shared due to ownership and licensing agreements with the FinnGen consortium. *Abbreviations: COPD, Chronic obstructive pulmonary disease*.

#### Outcomes

Outcomes were defined by the ICD-10-CM codes of the aforementioned T2IDs. For example, for AD, outcomes were PN (L28.1), BP (L12.0), VMR/AR (J30), CS (J32 or J33), asthma (J45), COPD (J44), EoE (K20.0) or food allergy (T78.0 or Z91.01). Any outcomes occurring prior to the index event were excluded during data retrieval.

#### Primary, sensitivity, and subgroup analyses

In the primary analysis, outcomes occurring any time from one day after the index event were considered. In sensitivity analysis S1, to exclude already present but nor diagnosed or coded outcomes at the index event, as well as to counter reverse causality, outcomes occurring any time from three months after the index event were considered. In S2, a minimal baseline time window of six months was mandated to ensure documentation of baseline characteristics and further mitigate detection bias. Like in the primary analysis, outcomes occurring any time from one day after the index event were considered. In S3, a stricter definition of the index event was used. In this analysis, a second documentation of the index event-defining criteria was required at least six months after the initial event. The subgroup analyses were stratified by age (1-12 years, ≥18 years), sex (female, male), and self-reported ancestry (Black or African American, Hispanic or Latino, or White).

#### Statistical analysis

A propensity score for each patient was generated by logistic regression analysis (with exposure as the dependent variable) using the Python package scikit-learn. Matching was performed 1:1 using the greedy nearest neighbor approach with a cut-off distance of 0.1 pooled standard deviations of the logit of the propensity score. Baseline characteristics were re-evaluated and reported after matching, differences were compared by t-test for continuous and z-test for binary or categorical variables. Relative risks and risk difference (RD) were calculated. Survival analysis was performed using the Kaplan-Meier method in Survival package v3.2-3 in R (R Foundation for Statistical Computing, Vienna, Austria) and validated by comparison with the outputs of SAS version 9.4 (SAS, Cary, NC). The proportionality assumption was tested by the coxph function in R’s Survival package. Kaplan-Meier--curves were compared using the Log-rank test. A univariate Cox proportional hazards regression was used to express hazard ratios (HR)s with 95%-confidence intervals (CI)s. To account for multiple comparisons, p-values were corrected using the Bonferroni method, such that p<0.0063 was considered statistically significant.

#### Genetic correlation studies

Genome-wide association summary statistics for seven major T2IDs (AD, asthma, PN, EoE, CS, COPD, AR and BP) were obtained from large publicly available datasets, including the FinnGen consortium, UK Biobank, GERA, and the Global Biobank Meta-Analysis Initiative (GBMI). For AD data were retrieved from a meta-analysis including 60,653 cases and 804,329 controls (25). For asthma: GWAS summary statistics were obtained from GBMI, comprising 121,940 cases and 1,254,131 controls from GERA (26). For VMR/AR summary statistics were obtained from both FinnGen (n=70,158) and GERA (27). Genetic data relating to COPD were drawn from GBMI (26). For CS: GWAS summary statistics were obtained from FinnGen (16,395 cases; n=299,737) and UK Biobank (3,026 cases; n=401,783). For EoE (28) with 1,930 cases and BP (FinnGen, **Table 1**) with 507 cases, no sufficiently powered GWAS summary statistics were available. Hence, existing GWAS for the two diseases were used. Food allergy was not included in the genetic correlation analysis because of lack of well-defined datasets. For asthma and AR, we conducted p-value weighted GWAS meta-analysis using METAL software. For each trait, we used GWAS summary statistics with European ancestry-based LD structure, and harmonized effect alleles and positions to GRCh37. Analyses were restricted to autosomal SNPs. All the GWAS summary statistics was aligned to 1000 genome project to correct for strand flips, allele mismatch and deviation from frequency from 1000 eur LD (> 20%). We next estimated pairwise genome-wide genetic correlations using LD Score Regression (LDSC), implemented in ldsc v1.0.1. The 1000 Genomes Phase 3 European reference panel was used to calculate LD scores. We applied a Bonferroni correction for multiple testing to define significance thresholds. Pairwise correlations with FDR-adjusted p-values < 0.05 were considered statistically significant. The presence of a significant genetic correlation (rg>0) was interpreted as evidence of shared heritability between traits.

#### Data sharing

All data relating to the retrospective cohort studies are included in the manuscript or the supplement tables. Data relating to the metanalyses and genetic correlations are available upon request from the corresponding author.

## Results

### Baseline characteristics

Data was sourced from a total of 129,702,895 EHRs. For the primary analysis, EHRs for patients diagnosed with incident T2IDs, specifically, CS (n=1,778,537), AD (n=774,536), food allergy (n=423,963), VMR/AR (n=3,487,860), PN (n=83,465), asthma (n=4,129,468), EoE (n=185,732), COPD (n=2,030,053), and BP (n=6,476), as well as between 13,108,725 and 13,421,991 for non-T2ID controls, were retrieved. Summarized baseline characteristics are found in **Table 1**, while detailed characteristics for all cohorts are presented in **Supplement Tables 2-10**. In all analyses, differences between groups observed prior to PSM were substantially reduced following matching.

### Chronic sinusitis is the core driver disease within the T2ID network

Across the disease progression network, chronic sinusitis emerged as the core driver disease risk within T2IDs. In the primary analysis, incident CS was significantly associated with increased risk of seven of eight investigated downstream T2IDs: Atopic dermatitis (HR 2.85, 95% CI 2.76-2.95, p<0.0001), PN (HR 2.34, 2.18-2.51), asthma (HR 3.12, 3.08-3.16), VMR/AR (HR 4.05, 4.01-4.10), COPD (HR 2.043, 2.007-2.079), EoE (HR 2.55, 2.36-2.75), and food allergy (HR 2.22, 2.15-2.30). These associations persisted across all three sensitivity analyses. In the primary and S1 analyses, an increased risk for BP was noted. This, however, was not validated in sensitivity analyses S2 and S3 (**Figures 2**–**3, Supplement Table 11**).

**Figure 2.**
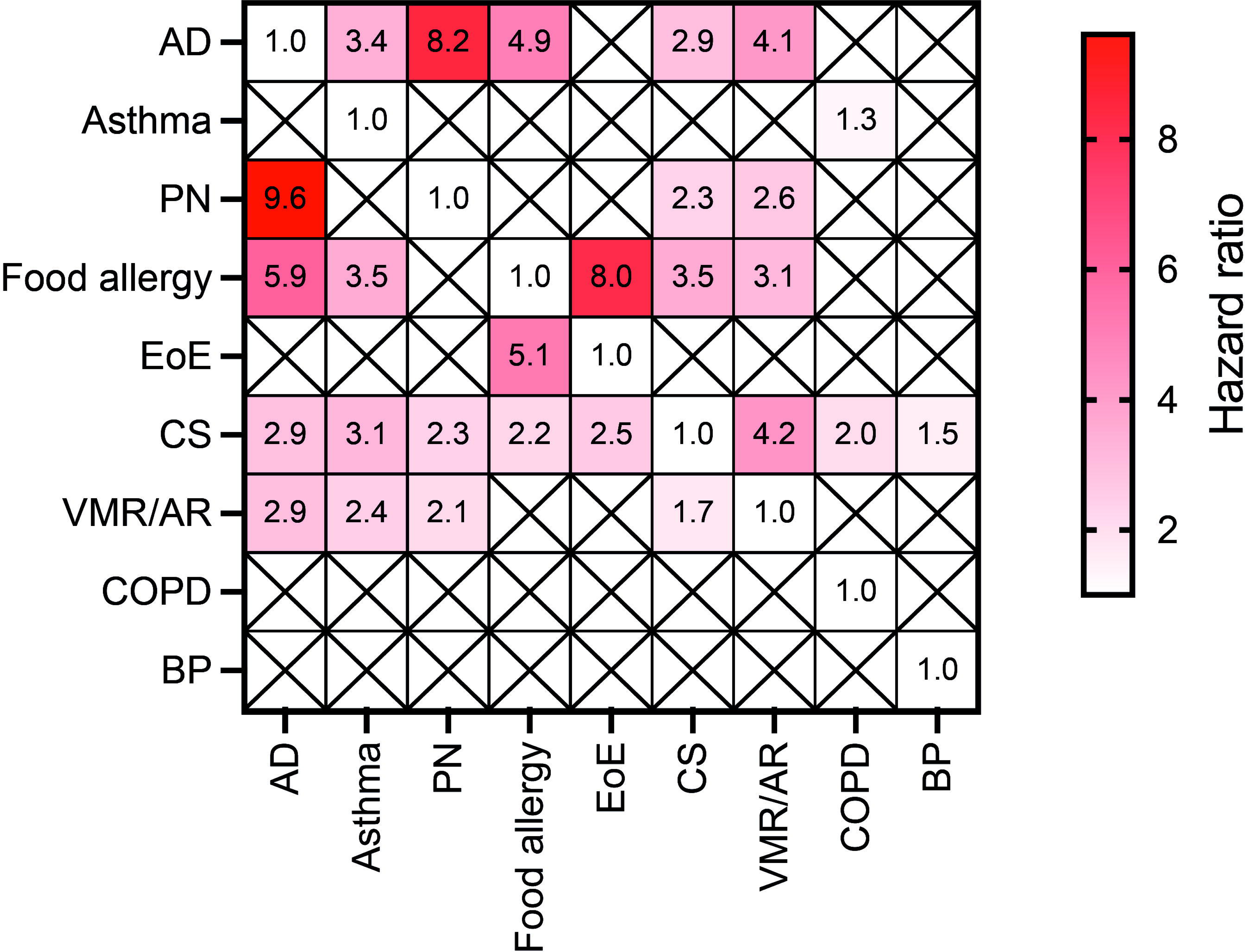
Cross-disease risk matrix of type 2 inflammatory diseases. Significant hazard ratios for incident type 2 inflammatory diseases (T2IDs) across all primary and sensitivity analyses (S1-S3). Each cell represents the HR from one T2ID (row) to another T2ID (column), calculated in longitudinal matched cohort analyses. Only statistically significant associations (α(adjust)=0.0063) are shown; nonsignificant comparisons are marked with an “X”. Analyses were conducted on the TriNetX electronic health record platform. Abbreviations: AD, atopic dermatitis; PN, prurigo nodularis; VMR/AR, vasomotor and allergic rhinitis; EoE, eosinophilic esophagitis; CS, chronic sinusitis; COPD, chronic obstructive pulmonary disease; BP, bullous pemphigoid.

**Figure 3.**
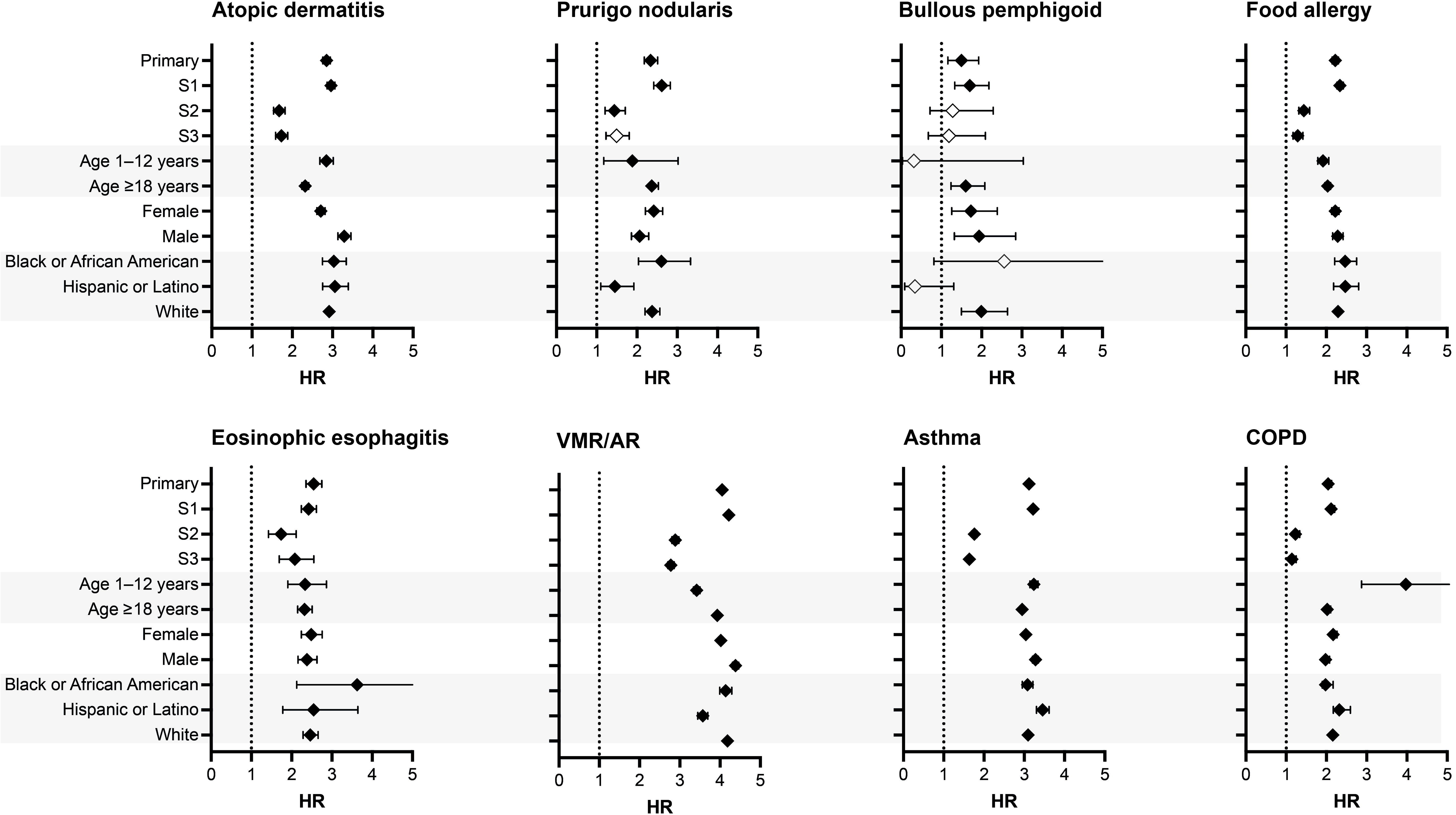
Risk of type 2 inflammatory diseases in patients with incident chronic sinusitis compared to non-type 2 inflammatory diseases controls. Hazard ratios (diamonds) and 95% confidence intervals (error bars) for the risk of developing other type 2 inflammatory diseases (T2IDs) in patients with chronic sinusitis (CS) versus non-T2ID controls. Each panel represents the indicated T2ID as an outcome, and each row shows results from the primary analysis, sensitivity analyses (S1-S3), and subgroup analyses stratified by age, sex, and race/ethnicity. Black diamonds indicate statistically significant results; open diamonds represent non-significant results. Alternating row shading is used to visually separate subgroup levels. Abbreviations: AD, atopic dermatitis; PN, prurigo nodularis; VMR/AR, vasomotor and allergic rhinitis; EoE, eosinophilic esophagitis; CS, chronic sinusitis; COPD, chronic obstructive pulmonary disease; BP, bullous pemphigoid.

**Figure 4.**
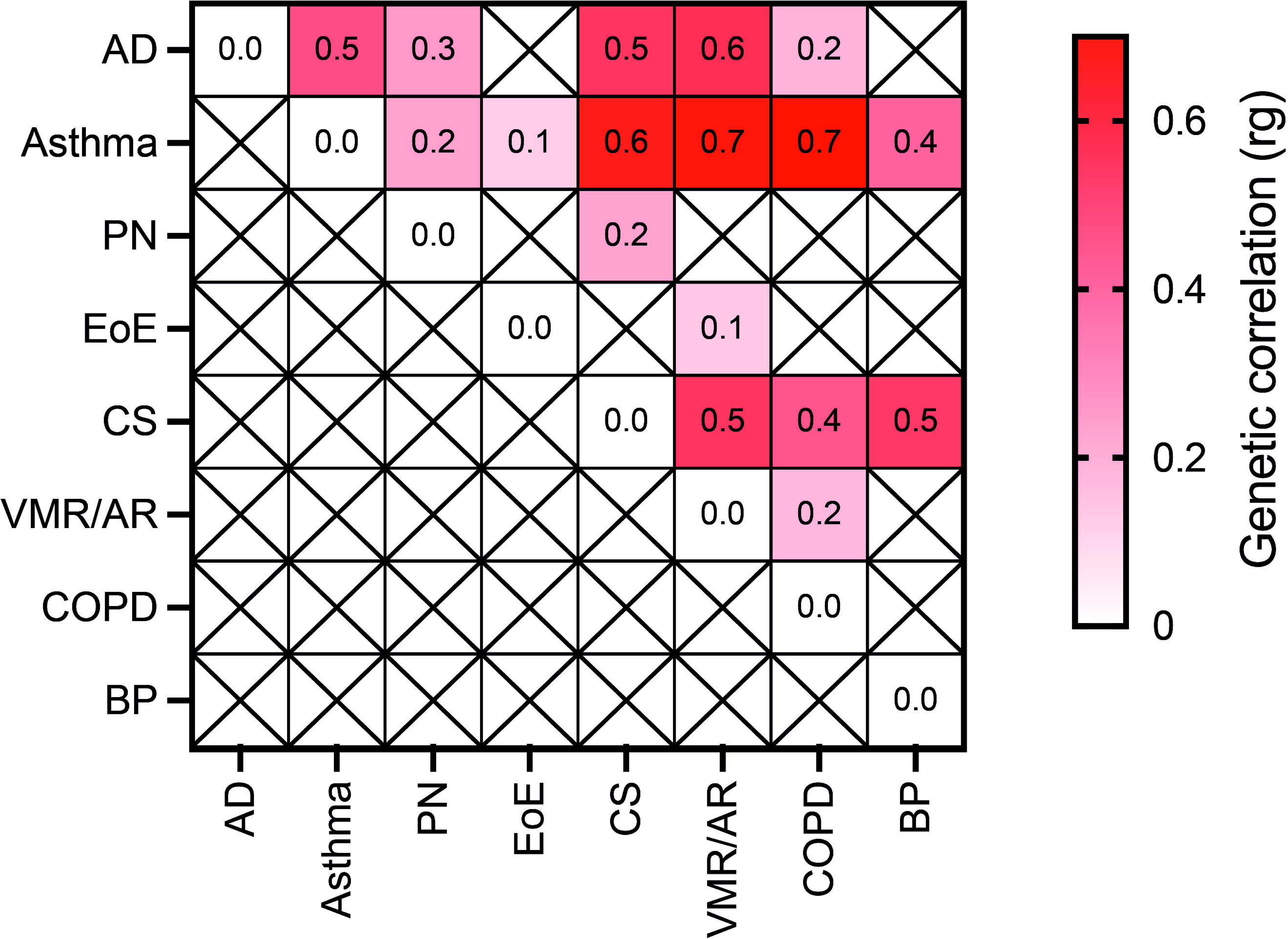
Genetic correlation matrix of type 2 inflammatory diseases. Pairwise genetic correlations (rg) between eight type 2 inflammatory diseases (T2IDs) estimated using LD Score Regression. Each cell indicates the magnitude of genetic correlation (rg) between the T2ID listed in the row and the T2ID listed in the column. Only statistically significant correlations (FDR-adjusted p□<□0.05) are shown; nonsignificant correlations are marked with an “X”. Color intensity reflects the strength of rg, with darker red indicating higher correlation. Genetic correlation analyses were performed using publicly available GWAS summary statistics and European ancestry LD reference panels. Abbreviations: AD, atopic dermatitis; PN, prurigo nodularis; VMR/AR, vasomotor and allergic rhinitis; EoE, eosinophilic esophagitis; CS, chronic sinusitis; COPD, chronic obstructive pulmonary disease; BP, bullous pemphigoid.

Subgroup analyses revealed significant differences among stratified groups: For PN, which typically affects the elderly (29), childhood CS did not alter the risk of subsequent PN diagnosis, whereas in adults, CS increased this risk (HR 2.36, 2.21-2.53). Conversely, the risk for a subsequent COPD diagnosis was lower in adult CS patients (HR 2.02) compared to children with CS (HR 3.98). By contrast, in the analyses stratified by either female of male risk, no impactful differences relating to the risk to be diagnosed with another T2ID following a CS diagnosis was noted. Racial disparities were noted for PN, where the highest risk increment imposed by CS was observed for those with Black of African American descent (HR 2.60), as opposed to White (HR 2.38) or Hispanic or Latino (HR 1.45) persons. Black of African American persons had also the highest risk for a subsequent EoE diagnosis (HR 3.63), compared to that of White (HR 2.46) or Hispanic or Latino (HR 2.55) persons (**Figure 3, Supplement Table 11**).

### Atopic dermatitis, food allergy and vasomotor and allergic rhinitis are relevant but lesser disease drivers among the T2ID network

AD also emerged as an important hub in the T2ID network, increasing the risk of five of the eight investigated T2IDs across the primary and all sensitivity analyses - specifically PN (HR 8.24, 7.12-9.55), VMR/AR (HR 4.16, 4.09-4.23), CS (HR 2.94, 2.85-3.03), asthma (HR 3.43, 3.37-3.50), and food allergy (HR 4.96, 4.77-5.15). In the primary analyses and in S1-2, AD was also associated with an increased risk for BP, EoE and COPD. However, in S3, this risk increment was not observed. Given the reciprocal risk increase between CS and AD, along with their high prevalence (30,31), both conditions should be recognized as major contributors to the development and perpetuation of T2IDs (**Figure 3**, **Supplement Table 12**). In the subgroup analyses stratified by age, major differences between childhood and adult incident AD patients were noted. Regarding PN, the risk increment imposed by AD amounted to a HR of 9.22 (7.88-10.80) in adult AD patients, which was, albeit still increased, not as pronounced in childhood AD (HR 6.33, 4.49-8.93). By contrast, the risks imposed for asthma and food allergy were comparatively higher in children with AD in contrast to adults (**Supplement Table 12**). When the analysis was stratified by sex, the risk imposed for other T2ID by AD was similar across both sexes, except for PN, with HR of 9.48 (7.62-11.79) in females and 8.39 (6.33-11.12) in males. Racial disparities were observed for the risks for PN (Hispanic or Latino < White < Black of African American) and food allergy (Black or African American < White < Hispanic or Latino) (**Supplement Table 12**).

Food allergy also increased the risk for the subsequent diagnosis of five of the eight investigated T2IDs, namely, AD (HR 5.94), VMR/AR (3.18), CS (2.50), asthma (3.50), and EoE (8.01). These risks were observed in the primary and all three sensitivity analyses. Compared to adult patients with food allergy, those aged 1-12 had a much higher risk association for the atopic dermatitis and EoE. In EoE, the HR in food allergy patients aged 1-12 years amounted to 9.82, while in those 18 years or older the HR for EoE was 5.39. In the subgroup analysis stratified by sex, no major differences in cross-T2ID risk were noted. When stratified by self-reported ancestry, racial disparities were noted. The risk of AD following a food allergy diagnosis was highest in those with Hispanic or Latino ethnicity (HR 6.10), as opposed to White (5.47) or Black or African American (5.14) individuals. Notably, for EoE, food allergy imposed the highest risk for EoE in Black or African Americans (8.67), while this risk increment was lower in Hispanic or Latinos (7.17) or Whites (6.10) (**Supplement Table 13)**.

Similarly, and across the primary and all sensitivity analyses, incident VMR/AR also increased the risk for four of the eight T2IDs investigated: AD, asthma, PN, and CS, with HRs ranging from 2.10 for PN to 2.90 for AD. Significant increased risks for food allergy, EoE, COPD, and BP were also noted in the primary and S1-2 analyses, but failed to replicate in S3. In the subgroup analysis no major differences relating to this risk were observed (**Supplement Table 14)**.

### Prurigo nodularis also contributes to cross-T2ID risk

PN consistently increased the risk for the subsequent diagnosis of AD (HR 9.62, 8.25-11.26), CS (HR 2.39, 2.22-2.58), and VMR/AR (HR 2.63, 2.49-2.78) in the primary and all sensitivity analyses. Diagnosis of PN did not change the risk for the other investigated T2IDs. In the subgroup analyses, adults, but not children with PN demonstrated an increased risk for AD, CS and VMR/AR. Furthermore, quite substantial racial disparities were noted: In persons with Hispanic or Latino ancestry, PN increased the risk for a subsequent AD diagnosis (HR 4.17). This risk for AD was further increased in those with White (HR 8.26) and especially Black or African American persons (HR 10.26). No major sex differences regarding to the risk of AD, CS, and VMR/AR were noted. Detailed information on these findings is presented in **Supplement Table 15**.

### Asthma, eosinophilic esophagitis, COPD, and bullous pemphigoid likely represent downstream manifestations of core T2IDs

Asthma increased the risk for a subsequent COPD diagnosis, which was most pronounced in children and in persons with self-reported Hispanic or Latino ethnicity (**Supplement Table 16**). EoE increased the risk of food allergy, which was post pronounced in children (**Supplement Table 17**). A diagnosis of COPD (**Supplement Table 18**) or BP (**Supplement Table 19**) was not associated with changes in the risk for any of the investigated T2IDs.

### Genetic correlation confirms the RWD-derived T2ID network and validates chronic sinusitis as a central T2ID driver disease

To validate and extend the findings from our retrospective cohort studies on cross-disease risks among T2ID, we next performed genetic correlation analyses. These were based on previously published GWAS meta-analyses as well as three newly conducted meta-analyses for AR, asthma, and CS. Detailed information on the non-FinnGen GWAS datasets is provided in **Supplementary Datasets 1-4**. In detail, we compared results from retrospective cohort analyses with genetic correlations for the 56 possible cross-disease comparisons. Among these, 34 combinations (61%) showed concordant signals, with both HR and rg being statistically significant and pointing in the same direction. In six of the comparisons (11%), a significant genetic correlation was observed, while only one direction of clinical association reached significance. This allows to provide a direction of the association of the two diseases when combining genetic correlation with retrospective cohort studies. In 16 of the comparisons (28%) the results were discordant. In 15 of these, a significant genetic correlation was observed in the absence of a significant HR in either direction. In two pairs, the inverse was true, with a significant HR but no detectable genetic correlation. Notably, most of these discordant significant genetic correlations were of modest magnitude. Together, these findings suggest that retrospective cohort studies may be more conservative in detecting disease associations compared to genetic correlation analyses, particularly when genetic overlap is limited (**Figure 5**).

**Figure 5.**
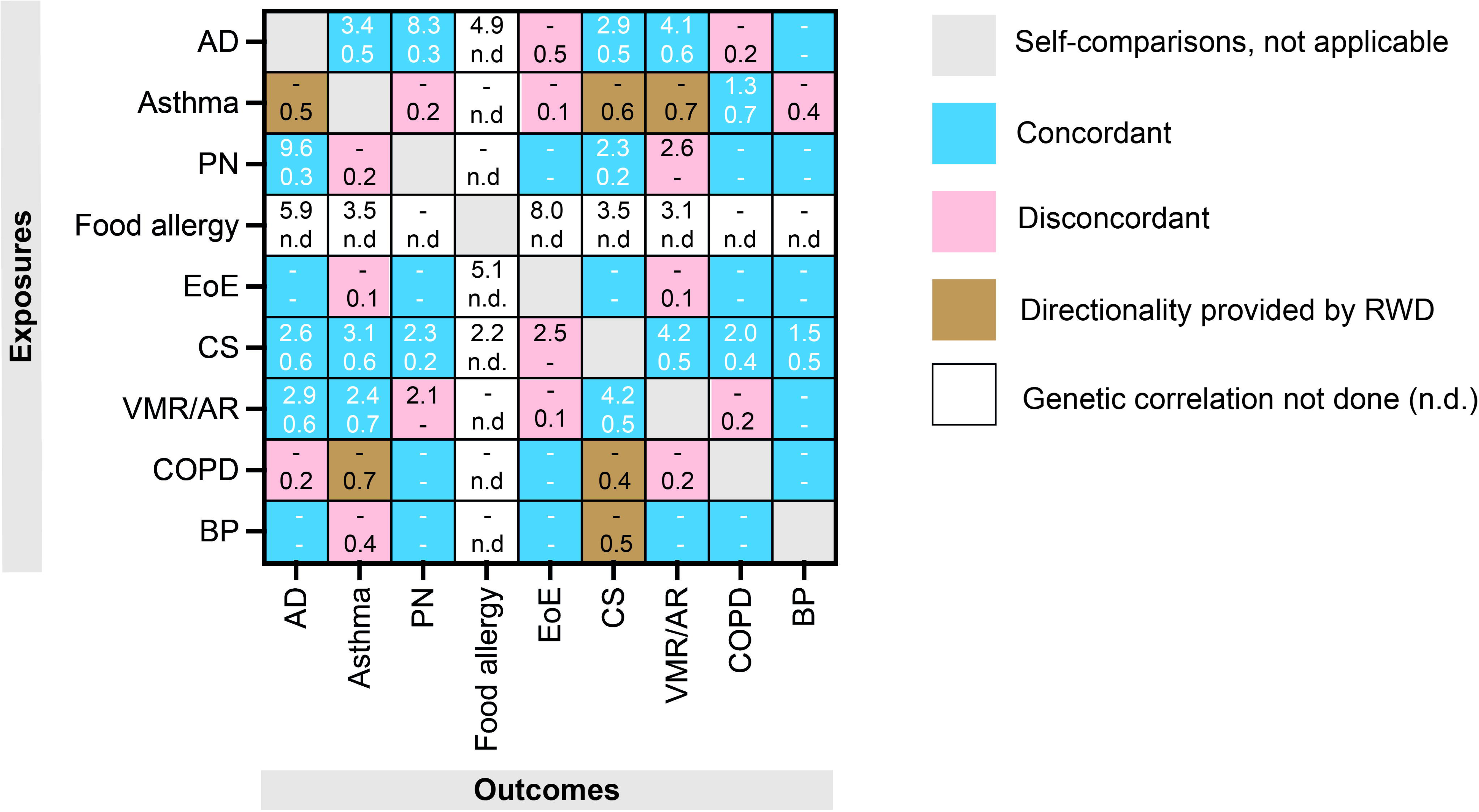
Cross-modal matrix of real-world and genetic correlations among type 2 inflammatory diseases. Each cell represents a disease pair, with the top number indicating the hazard ratio (HR) for the real-world risk of developing the outcome disease (based on retrospective electronic health record data) and the bottom number indicating the genetic correlation (rg) derived from LD Score Regression. Color coding reflects the concordance of the associations: Blue indicates concordant associations where both HR and rg are significant and directionally consistent; pink indicates discordant associations; brown indicates that directionality is provided by real-world data; and white indicates that genetic correlation analysis was not performed due to lack of GWAS of well-defined cohorts. Gray cells indicate self-comparisons. Only statistically significant associations are shown (p<0.0063 for HRs; false discovery rate–adjusted p<0.05 for rg). Non-significant comparisons are marked with a dash (-), and missing genetic data are denoted as “n.d.” Abbreviations: AD, atopic dermatitis; asthma; BP, bullous pemphigoid; COPD, chronic obstructive pulmonary disease; CS, chronic sinusitis; EoE, eosinophilic esophagitis; PN, prurigo nodularis; VMR/AR, vasomotor and allergic rhinitis.

We next examined chronic sinusitis in more detail, given its central role in driving the T2ID comorbidity network. To continue with chronic sinusitis, we observed that genetic correlations aligned well with the increased risks identified in the retrospective cohort data for several diseases, including asthma, AD, PN, VMR/AR, COPD, and BP. Notably, RWD also revealed an increased risk of EoE in patients with chronic sinusitis, which was not reflected in the genetic correlation analysis. In several instances, the combination of RWD and genetic correlation enabled directional interpretation of the associations. For example, CS and asthma share a strong genetic correlation (rg=0.6), and RWD showed that patients with CS have an increased risk of subsequently developing asthma (HR 3.12, 3.087-3.16). In contrast, asthma does not confer a significantly increased risk for CS (HR 1.30, 1.29-1.32). This directional asymmetry suggests that the shared genetic architecture between CS and asthma predominantly contributes to asthma development in individuals with CS, rather than vice versa.

## Discussion

In this study, we used two independent and complementary approaches, namely retrospective cohort analyses from a large-scale EHR database and genome-wide genetic correlation estimates, to systematically map comorbidity patterns across nine major T2IDs. Our findings provide an in-depth description of the interrelated network of disease trajectories amongst T2IDs. This network-based view goes far beyond the classical concept of the atopic march, which describes a unidirectional sequence from AD to asthma and AR (11). Instead, our data reveal that the atopic march represents just one of many possible comorbidity trajectories within a broader, more heterogeneous spectrum of T2ID interactions that span multiple organ systems and diverse disease pairings. Within this network, CS emerged as a central hub, significantly increasing the risk for nearly all other T2IDs examined. AD, food allergy, and VMR/AR also contributed to network propagation, albeit with smaller impact. These observations remained robust across multiple sensitivity analyses and diverse patient subgroups.

The traditional concept of the atopic march describes a sequential progression of T2IDs, starting with AD in infancy, followed by food allergies, AR, and asthma. This model has been instrumental in understanding the development of T2IDs (11). However, emerging research suggests that this linear trajectory does not encompass the full complexity of T2ID progression (16,32,33). Furthermore, the concept of the atopic march exclusively focuses on disease trajectories in childhood, with T2ID comorbidity patterns in adulthood being less clear (33). Our results contribute to this evolving understanding, demonstrating a complex and intricate network of cross-disease trajectories among T2IDs, extending beyond the traditional atopic march. Notably, our data identify CS as a central driver within the T2ID network, significantly increasing the risk for multiple downstream T2IDs. This observation challenges the conventional view that positions AD as the primary initiator of the T2ID comorbidity risk.

Importantly, our study incorporates stratified analyses by age, sex, and race/ethnicity, revealing marked disparities in disease trajectories. For example, adults with CS or AD had substantially higher risks for developing PN or COPD, while children with food allergy showed an elevated risk for developing EoE. Racial disparities were also pronounced: Black or African American individuals, compared to White or Hispanic/Latino individuals, often displayed the highest cross-disease risks, particularly for PN and EoE, compared to other racial groups. These findings align with a higher PN incidence in Black or African American individuals compared to Whites (34), but contrasts findings in EoE that has an overrepresentation of White individuals (35). This discrepancy may suggest that racial disparities in T2IDs differ between incident disease risk and subsequent disease manifestation within comorbidity trajectories. These findings reinforce the need to consider patient history as well as demographic context when evaluating disease progression and risk.

To determine whether observed comorbidities reflect shared etiologic mechanisms or simply co-occurrence, we complemented RWD analyses with genetic correlation studies. Across 56 possible disease pairs, the majority showed concordant associations in both data modalities, supporting shared heritability and potential biological connections. In a few cases, only one clinical direction was significant, but genetic correlation provided evidence of shared architecture, enabling directional inference. Some comparisons yielded discordant findings. Most of these involved a significant genetic correlation in the absence of a clinical signal, suggesting that genetic methods may be more sensitive to low-penetrance or latent disease relationships. Alternatively, these observed genetic correlations without RWD validation may reflect a shared genetic architecture due to pleiotropy, co-inheritance, or polygenic overlap, but without any mechanistic links amongst the two diseases.

Our study has several key strengths. First, it leverages two independent and complementary methodological approaches: large-scale, RWD from over 129 million individuals and genome-wide genetic correlation analyses. Second, the findings were validated across multiple sensitivity analyses and stratified by age, sex, and race/ethnicity, allowing for robust assessment of disparities and generalizability. Third, the integration of genetic correlation enables insights into shared heritability and helps infer potential directionality where observational data alone may be ambiguous. Nonetheless, several limitations should be considered when interpreting these findings: First, the study retrospectively analyzed RWD and is therefore subject to potential bias from unmeasured confounders. A key concern is the definition of incident T2IDs, as incident diagnoses are based on the first documentation at a participating HCO within the TriNetX network. This approach may fail to capture prior diagnoses made outside the network. In addition, RWD sources are susceptible to miscoding of ICD codes and other inaccuracies. Healthcare access disparities may also influence the documentation of comorbidities, potentially masking measured socioeconomic differences. Finally, while our findings demonstrate consistent associations between specific T2IDs, both clinically and genetically, they do not imply causality. Regarding the genetic correlation analysis, the limited sample sizes for BP EoE likely resulted in reduced heritability estimates and diminished power to detect significant genetic correlations. Although temporal sequencing in RWD and concordant genetic correlations increase the biological plausibility of certain disease transitions (e.g., CS to asthma), genetic correlation reflects shared heritability rather than direct causation. Residual confounding cannot be fully excluded. Future studies using Mendelian randomization, longitudinal designs, and mechanistic experimentation are needed to determine causal relationships more definitively.

Our findings underscore that T2IDs form a complex and tightly interwoven network of cross-disease trajectories. Within this network, CS is a key upstream driver of downstream T2ID comorbidities. CS, and to a lesser extent also AD, food allergy and VMR/AR, may serve as early indicators (and actionable targets) for preventive intervention to halt emerge of further T2ID. Crucially, the identified sex- and race-specific disparities in comorbidity patterns highlight the need for personalized risk stratification and more equitable, precision-driven approaches in patients diagnosed with incident T2ID.

## Supporting information

Strobe Checklist

## Acknowledgements

We express our gratitude for the exceptional and ongoing support provided by Friederike Uebing (Justiziariat und Stabsstelle Vernetzung und Strategische Kooperation, UKSH, Campus Kiel) in facilitating access to the TriNetX platform at the UKSH.

